# Natural history of a parasite–induced biliary cancer

**DOI:** 10.1101/2024.09.20.24313287

**Authors:** Thomas Crellen, Francesca Vita, Chiara Braconi, Paiboon Sithithaworn, T. Déirdre Hollingsworth

## Abstract

Pathogens are major drivers of cancer globally and the processes of infection and carcinogenesis unfold over decades making them difficult to observe in human or natural populations. We investigate these hidden dynamics for the foodborne trematode *Opisthorchis viverrini*, which is a primary cause of biliary cancer (cholangiocarcinoma) and infects 12 million people in Southeast Asia. In tumors from patients exposed to *O. viverrini* we find that the earliest chromosomal amplifications carrying driver genes occurred at 30 years old on average, two to four decades before cancer diagnosis, and disproportionately contain *TP53, PTEN* and *FGFR2* genes. We then fitted transmission models to parasitological data from Thailand spanning 27 years (*n* = 11,517) finding that, for people born between 1960–1989, first exposure occurred at two years old and by 30 years individuals had been cumulatively infected with a median of 72 worms. Trematodes are long-lived and our analysis quantifies the average lifespan of *O. viverrini* as 13 years (90% credible interval [CrI] 7–26 years) within human hosts. The lifetime probability of diagnosis with cholangiocarcinoma is 4.9% (90% CrI 4.7–5.0%) given prior exposure to *O. viverrini*, which is fourteen-fold higher than in populations non-endemic for the parasite. We find strong evidence for a dramatic decline in parasite transmission from 1990 onwards in Thailand, suggesting that the incidence of cholangiocarcinoma will decline over the coming decades. Our study is the first to demonstrate how pathogen exposure drives patterns of cancer within a population and provides evidence for therapeutic and public health interventions.

## Introduction

Infectious organisms are a major contributor to the burden of human cancers globally (1), thus challenging the classic distinction between ‘communicable’ and ‘non-communicable’ diseases. The epidemiology of pathogen–induced cancers includes the transmission of the infectious agent, which is a dynamic population–level process, plus the resulting within– host pathology that drives carcinogenesis (2). These processes are complex, unfold over many years, and are unlikely to be co-observed in a single prospective cohort (3).

Of the eleven pathogens recognised as direct human carcinogens; three are parasitic trematodes^*†^. Infection with the foodborne liver flukes *Opisthorchis viverrini* and *Clonorchis sinensis* cause biliary cancer (cholangiocarcinoma), whilst the waterborne blood fluke *Schistosoma haematobium* causes cancer of the bladder (squamous cell carcinoma). The pathology arising from infection with parasitic worms is typically chronic as definitive hosts rarely develop protective immunity to reinfection, making it difficult to quantify the impact of any single helminth species over decades of exposure and in populations with co–infections (4, 5).

Our study focuses on the liver fluke *O. viverrini*, which is acquired by eating raw freshwater fish; a traditional component of the diet in regions of Southeast Asia (6). The fluke has a complex lifecycle, which involves asexual reproduction within freshwater snails and encysts as a mammalian–infective stage (metacercariae) in cyprinid fish. Despite public health programs to control the parasite, an estimated 12 million people were infected with *O. viverrini* across Thailand, Lao PDR, Cambodia and Vietnam in 2018 (7). While the prevalence has shown gradual declines in Thailand due to parasite control programs during the second half of the twentieth century (8), progress has recently slowed (9, 10). In Cambodia and Lao PDR, by contrast, there is evidence of increased transmission over the past two decades (7, 11). Liver fluke–endemic countries have the highest incidence of cholangiocarcinoma globally and cases of hepatic cancers in these regions are disproportionately attributable to cholangiocarcinoma, rather than hepatocellular carcinoma (12, 13). The mechanisms through which flukes induce cholangiocarcinoma is a combination of mechanical damage, inflammation of the biliary epithelium, and the secretion of proteins; in particular the peptide granulin (14).

Given the poor prognosis for cholangiocarcinoma (15) and the preventable nature of parasite infection, there is a strong motivation to understand the link between liver fluke exposure and carcinogensis in humans (16). Prior to the onset of driver mutations, anthelmintic treatment and reducing parasite exposure should be prioritized as public health interventions, whereas after the onset of irreversible malignancies the priority for interventions shifts to ultrasound screening for liver pathology and early referral for hepatobiliary surgery (17).

This study infers the timings of driver mutations for fluke– induced biliary cancer using computational methods that characterize the evolution of tumors from a single biopsy (18, 19). We then define the age of first exposure to the parasite by fitting dynamic transmission models to parasitological survey data from Thailand. Finally, we estimate the lifetime probability of diagnosis with cholangiocarcinoma given infection with *O. viverrini*. By combining evolutionary cancer genomics with epidemiological analysis, we obtain unique insights into the relationship between pathogen exposure and tumorigenesis, with implications for evidence–based disease control.

## Results

### Cholangiocarcinoma tumor genomes

We obtained paired tumor and normal whole–genome sequences from cholangiocarcinoma patients who were previously infected with liver fluke and treated at a large public hospital in Northeast Thailand (20). We used a bioinformatics pipeline to call somatic single nucleotide variants (SNVs), copy number alterations, and inferred the clonal status of these mutations (see Methods). The age of the patients at surgery ranged from 37–79 years (median 57 years), 50% of patients were female, and all were born prior to 1980 (20). After mapping reads, variant calling, and filtering (see Methods) we obtained 2,349–27,821 (median 10,360) SNVs and 268–14,230 (median 1,382) somatic indels per tumor. The overall ploidy (chromosomal copies) per tumor ranged from 1.2–3.7 (median 2.0).

### Evolution of cholangiocarcinoma tumors

We estimated the timing of driver mutations in 43 genes implicated in cholangiocarcinoma development (22, 23) using evolutionary models that time chromosomal amplifications using the number of accumulated SNVs on different chromosomal copies. We first assessed whether any tumors had been subjected to whole– genome duplication events based on the correlated timings of chromosomal amplifications throughout the genome (19) and concluded that this had occurred in three tumors (see Fig. 1A). Across all tumors, the copy number was disproportionately higher in chromosomes 7 and 17. In 17/22 tumors with sufficient ploidy and tumor purity, we inferred the timing of focal chromosomal amplifications in potential driver genes for cholangiocarcinoma. Overall the majority of amplifications (166/287; 58%) occurred later in ‘chronological time’ (0.75 or later), with a smaller proportion (46/287; 16%) occurring earlier in tumorigenesis (before 0.5), as shown in Fig. 1B.

**Fig. 1.**
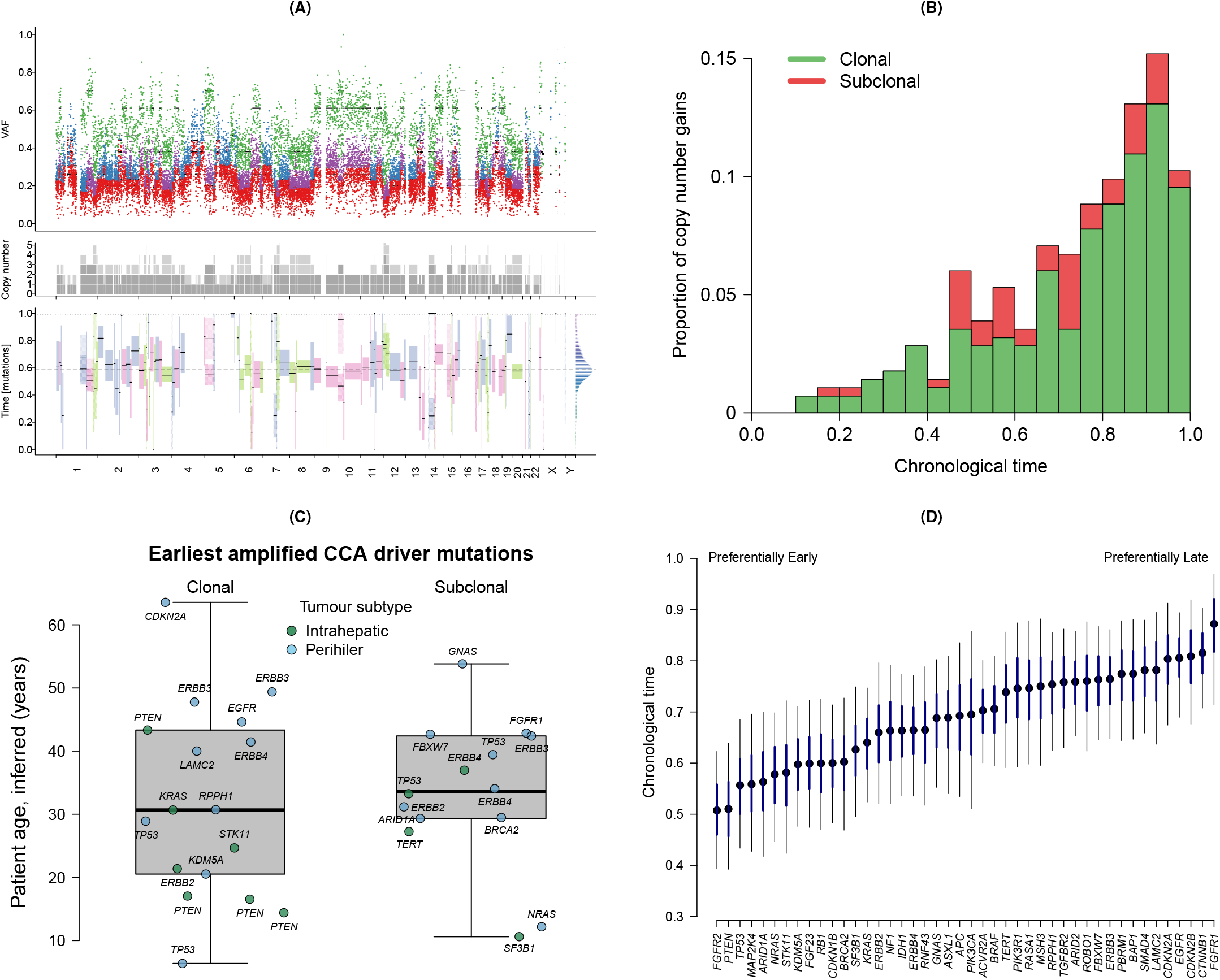
Evolution of cholangiocarcinoma tumors. A) Subclonal lineage reconstruction for cholangiocarcinoma tumor CCA_TH19 (19). The upper plot shows the variant allele frequency (VAF) for SNVs which are colored by clonal state; clonal (early) = green, clonal (late) = purple, clonal (unknown) = blue, subclonal = red. The middle plot shows the inferred copy number frequency (ploidy) by chromosome. The lower plot shows the inferred timings of copy number gains. For this tumor, the timings of copy number gains are correlated across chromosomes, indicating a whole-genome duplication event at a ‘mutation time’ of 0.6. B) Histogram showing the timing of 287 clonal and subclonal amplification events for 44 driver mutations in 17 cholangiocarcinoma tumors (21). C) The earliest amplified clonal and subclonal driver mutations in cholangiocarcinoma (CCA) tumors, where the timing is estimated using CpG>TpG mutations multiplied by the patient age at surgery (20) to give the inferred age. Each point represents the first amplified driver gene per patient, labeled with the gene name, and the points are colored by tumor anatomical subtype (intrahepatic or perihilar). The overlaid box and whisker plots shows the median and interquartile range. D) Estimates of the chronological time of amplification by gene. Results are shown from a generalized linear model applied to estimates from AmplificationTimeR (21). The black points give the posterior median, the thick blue line gives the 50% credible interval and the thin black line the 95% credible interval.

### Timing of amplified driver mutations

We determined the earliest amplified driver gene for each tumor sample, and classified these as either ‘clonal’ (occurring on the most recent common cell lineage of the tumor) or ‘subclonal’ (a subsequent clonal expansion within the tumor that has not risen to fixation) (24). To calculate the age at which these amplifications occurred in patients, we scaled the chronological time estimate with the patient’s age at surgery (see Methods). The first clonal amplification of driver genes occurred at a median age of 30 years and with an interquartile range (IQR) of 20–43 years (Fig. 1C). The first subclonal amplifications occurred at a median age of 33 years (IQR 29–41 years). A variety of genes were the earliest amplified, with the most frequently occurring being the tumor suppressor *PTEN* in the clonal lineages (earliest in four tumors and amplified in 7/17 tumors), while the tumor suppressor *TP53* and the protein kinase *ERBB4* were the most common in subclonal lineages (earliest in two tumors and amplified in 7/17 and 6/17 tumors respectively) see Fig. 1B. Our timing of somatic events uses C>T mutations at CpG sites (CpG>TpG; also known as single base substitution signature 1 [SBS1]) which has clock–like properties as the mutational load correlates with age in cancer patients (25). However, the CpG>TpG mutation rate may accelerate with age in cancerous cells (19). We therefore compared the findings above from a constant rate model against alternative scenarios where the CpG>TpG mutation rate accelerates 2, 5, 8 or 12–fold during tumorigenesis. In a scenario where the CpG>TpG mutation rate accelerates 5–fold over the lifetime of the patient, the age of earliest amplified driver genes for cholangiocarcinoma increases to 36 years on average (IQR 28–46 years).

To determine whether certain genes are disproportionately likely to be amplified early or late within the lifespan of the tumor, we applied a generalized linear model to the chronological age estimates for each of the amplification events, while controlling for host sex (female [reference] or male), tumor anatomical subtype (intrahepatic [reference] or perihilar) and clonality (clonal [reference] or subclonal), see Methods and Equation 2. We restricted this analysis to amplification events with at least ten CpG>TpG SNVs, giving 271 amplification events from 17 tumors. The model estimates per gene are shown by chronological age in Fig. 1D. Overall, the driver genes which were disproportionately found to be amplified early were the fibroblast growth factor receptor *FGFR2, PTEN* and *TP53* (these genes were amplified in 7 tumors), while the tumor suppressor genes *BAP1* and *PBRM1* (both amplified in 5 tumors), and the receptor gene *FGFR1* (amplified in 4 tumors) were amplified later in tumorigenesis. The coefficients from the generalized linear model indicate that clonality did not affect gene amplification times (odds ratio [OR] 0.96; 0.74–1.27), although male patient sex (OR 1.23; 90% CrI 1.00–1.55) and perihilar anatomical subtype (OR 1.39; 90% CrI 1.12–1.71) were both associated with later timings of amplification events. Our findings add support to previous studies that have noted early clonal amplification of *TP53*, in particular, as a driver across a range of cancer types (19).

### Liver fluke transmission

As fluke–induced pathology of the biliary tract is chronic, to understand the etiology of cholangiocarcinoma it is necessary to consider prior exposure to the parasite at the individual or population level which can be estimated from historical parasitological surveys (2). Therefore, we collated epidemiological surveys from Thailand in which diagnostic observations of *O. viverrini* infection intensity (worm burdens or fecal egg counts) were available by host age. Our analysis uses data from 4,056 individuals obtained from seven surveys conducted between 1980–1989, prior to the onset of large-scale control programs against liver fluke (8), and 7,448 individuals from five surveys conducted between 1994–2017, which are summarized in Table 1. We fitted a mechanistic parasite transmission model to these data where the parasite burden at a given age is the result of flukes infecting the host with an age-variable transmission rate, referred to as the ‘force of infection’, and flukes exiting the host due to the spontaneous death rate of adult worms (26). See Methods for model details.

**Table 1.**
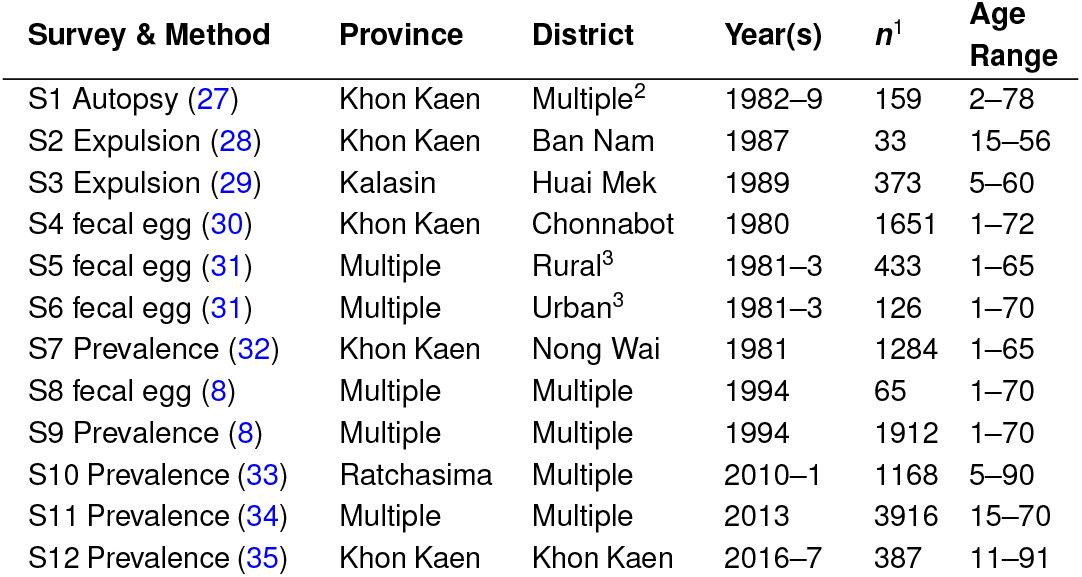
Cross–sectional surveys analyzed in this study investigating the relationship between host age and *Opisthorchis viverrini* worm burden, fecal egg counts, or prevalence by fecal egg diagnostic (Fig. 2A). Surveys were conducted in Northeast (N.E.) Thailand either before the onset of liver fluke control programs (S1–S7; pre–intervention), or following a national control program in the early 1990s (S8–S12; post–intervention). Age range of the participants is shown in years. ^1^Sample size of human participants in survey. ^2^Autopsy cases were from N.E. Thailand, with the majority from Khon Kaen province. ^3^Hospital–based study where patients from N.E. Thailand were recruited and classified as originating from either rural or urban communities.

### Exposure to liver fluke in human populations

The average age at which a person born between 1960–1989 was first infected with a single *O. viverrini* fluke is 2.2 years old (90% prediction interval [PI] 1.8–3.3 years). The average age of first infection for a person born after 1990 increases to 8.5 years (90% PI 7.5–10.5 years), although a smaller proportion of the population ultimately becomes exposed during their lifetime; 32% post–intervention (90% CrI 26–39%) compared with 88% pre–intervention (90% CrI 83–91%). We define exposure to *O. viverrini* as the cumulative number of adult flukes acquired by a person over time, which is given by the area under the force of infection curve (Equation 6). In endemic regions of Thailand in the 1980s by ages 10, 20 and 30 years the median number of *O. viverrini* cumulative flukes was 12 (90% PI 8–16), 39 (90% PI 27–52) and 72 (90% PI 48–103) respectively (Fig. 2C). Following public health interventions in the 1990s (8), we estimate that the *O. viverrini* force of infection declined substantially, with an almost 40–fold reduction in the age–dependent transmission rate, resulting in the majority of individuals born after 1990 remaining uninfected at age 30 (median worm burden is zero). The distribution of *O. viverrini* parasites among human hosts shows high variation, as is characteristic for helminths (36), and for the most heavily infected decile the cumulative burden is 691 worms by 30 years of age in the pre–intervention period (90% PI 504–986) and 14 worms in the post–intervention period (90% PI 10–21). These findings add weight to the reported declines in *O. viverrini* transmission in Thailand following control programs (7–9), though our analysis is the first to quantify this effect in terms of worm burden, age of first exposure, and proportion of the population exposed.

**Fig. 2.**
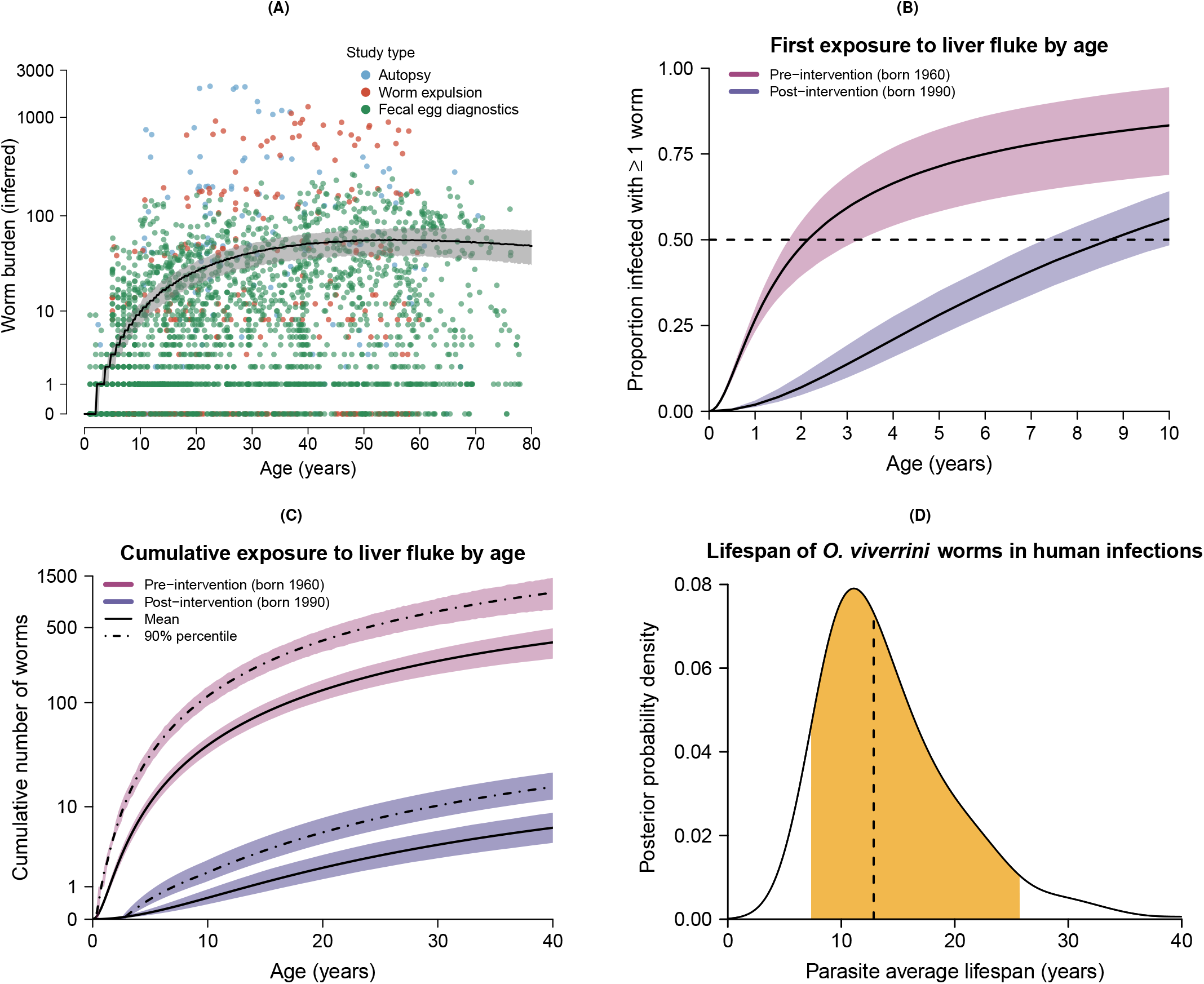
Epidemiological insights into the carcinogenic liver fluke *Opisthorchis viverrini* in Thailand. A) Worm burdens of *O. viverrini* by participant age, inferred by a parasite transmission model from multiple surveys conducted between 1980–1989 prior to large–scale interventions. The data used for model fitting were counts of i) adult worms obtained from autopsy, or ii) worm expulsion, or iii) parasite eggs in faeces (data shown from S1–S6, see Table 1). Observed data were transformed into inferred worm burdens using Equations 3, 8, 14; see Methods and (37). The fitted black line shows the simulated median worm burden by host age, with the shaded area giving the 90% prediction interval (model fitted to data from S1–S7; see Table 1). The y–axis is log–transformed. B) Proportion of exposed population with at least one parasite in the pre–intervention (1980–1989; S1–S7) and post–intervention periods (1994–2017; S8–S12 see Table 1), as inferred by the parasite transmission model. Solid lines are central estimates and shaded areas give the 90% credible interval (CrI). Where the solid lines cross the dashed line, this gives the age at which half of the exposed population become first infected; 2.2 years old (90% CrI 1.8–3.3 years) in the pre–intervention period and 8.5 years old (90% CrI 7.5–10.5 years) in the post–intervention period. C) Cumulative exposure to *O. viverrini* by age (Equation 6). The solid lines show the mean for the pre–intervention and post–intervention periods, while the dot–dash lines show the cumulative worm burden for the upper 90% population percentile. Shaded areas give the 90% prediction interval. The y–axis is log–transformed. D) Average lifespan of adult *O. viverrini* worms in human hosts. The posterior probability density is shown here for *log*(2)*/σ*; where *σ* is the spontaneous death rate of adult worms (Equation 3). Dashed line gives the posterior median (13 years). The filled area gives the 90% CrI (7–26 years).

### Lifespan of the adult worm

The lifespan of helminths in human infections is considered to be in the order of one year to three decades, although this is rarely quantified (38). We estimate the lifespan of *O. viverrini* within our force of infection model as 12.9 years (90% CrI 6.2–23.1 years), which is defined as the average time for half of adult stage parasites to die within the human host in the absence of anthelmintic treatment (Equation 3, see Methods). Our posterior distribution for parasite mortality is long–tailed (Fig. 2D) with the top five percent of adult *O. viverrini* lifespans ≥ 25.7 years. This finding is consistent with a case report of the related liver fluke *C. sinensis*, which persisted in an emigrant for 26 years (39). We note that the posterior distribution of the worm mortality parameter is correlated with a force of infection parameter and therefore encourage the use of the interval for *O. viverrini* lifespan (7–26 years), rather than the point estimate, in future analyses.

### Latent and induction periods of cholangiocarcinoma

Bringing together the evidence presented thus far, we hypothesize that people born 1960–1989 in Northeast Thailand first became infected with liver fluke in early childhood and driver mutations for biliary cancer occurred around three decades later. We sought to validate our hypothesis using cancer registry data consisting of 10,737 cases of cholangiocarcinoma diagnosed in Northeast Thailand between 1985–2009 (40). The median age of diagnosis was 59 years between 1985–1997 and 63 years between 1997–2009. Using a time-to-event analysis, which accounts for interval censored data, we estimate the age of driver mutation and the subsequent age of cholangiocarcinoma diagnosis as sequential gamma distributions (see Methods). The induction period (time from initial parasite infection to driver mutation) is estimated as 28 years (90% CrI 16–39 years) and the latent period (time from driver mutation to cancer diagnosis) is 32 years (90% CrI 21–44 years). These distributions are shown in Fig. 3A. The cancer incidence data therefore support our estimate of the time to first amplified mutation at around thirty years of age, given exposure by age two. The estimated latent period from registry data is longer than the 22 years indicated by our analysis of tumor genomes (time from first amplified driver mutation to biliary surgery), which likely reflects an older average age of diagnosis in the registry data compared to our much smaller sample of patients with sequenced biliary tumors. Previous research on the induction and latent periods for fluke–induced cholangiocarcinoma in humans is limited, though American veterans from the Vietnam War had an elevated incidence of biliary cancer five or more decades after potential exposure to *O. viverrini* between 1965–1971 (41). While the association between military service in Vietnam and fluke–induced cholangiocarcinoma is controversial (42), the reported time between parasite exposure and cancer is consistent with our findings.

**Fig. 3.**
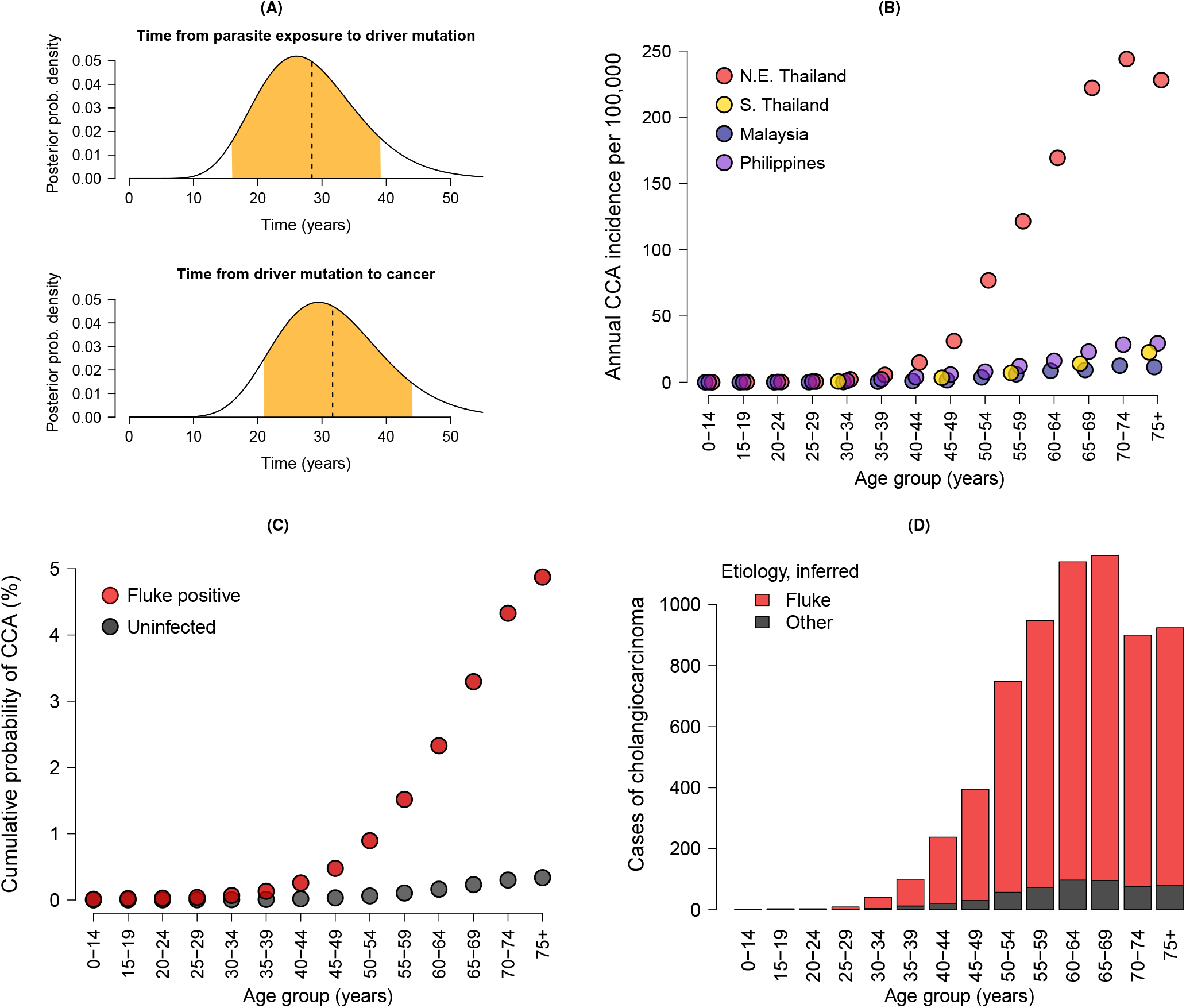
Incidence and age–distribution of cholangiocarcinoma (CCA) at the population level. A) Time–to–event distributions for the induction period (parasite exposure to driver mutation; median 28 years) and the latent period (driver mutation to cancer diagnosis; median 32 years). The plots show posterior probability distributions, where the dashed line gives the median and the orange area the 90% credible interval (CrI). B) Annual incidence of cholangiocarcinoma per 100,000 people by age group for Northeast (N.E.) Thailand, where the carcinogenic liver fluke *Opisthorchis viverrini* is endemic, and Southern (S.) Thailand, Malaysia and the Philippines, which are non-endemic for *O. viverrini*. C) Cumulative lifetime probability of diagnosis with cholangiocarcinoma in Southeast Asia for i) a person infected with liver fluke (dark red) and ii) an uninfected person (grey). The model output is shown from a survival analysis (Equation 16, see Methods). Points give the posterior median probability within each age group. D) The age distribution of 10,737 cases of cholangiocarcinoma from Northeast Thailand between 1985–2009 (40), colored by etiology (fluke or other) inferred from the survival model posterior parameters.

### Lifetime probability of cholangiocarcinoma

Finally, we estimate the lifetime probability of acquiring cholangiocarcinoma, and the increase in risk from parasite infection, by fitting a survival model (see Methods and Equations 16 and 17) to age-incidence cancer registry data from Northeast Thailand between 1997–2009 (40). For comparison, we include cancer registry and mortality data from neighboring populations (Southern Thailand, Malaysia and the Philippines) that are non-endemic for *O. viverrini* to estimate the baseline risk of cholangiocarcinoma in the absence of parasite infection. The underlying data are shown in Fig. 3B as the annual incidence of cholangiocarcinoma per 100,000 people by age. We obtain posterior parameters for age–variable intercepts (*α*_*a*_) and a coefficient (*β*), which gives the relative risk of diagnosis with cholangiocarcinoma given exposure to liver fluke, estimated as 2.73 on the logit scale; equating to an odds ratio of 15.3 (90% CrI 14.7–15.8).

We use the posterior parameters to produce counterfactual scenarios for the lifetime probability of cholangiocarcinoma diagnosis given prior infection with liver fluke, see Fig. 3C. Our results show that by 75 years of age the cumulative probability of diagnosis with cholangiocarcinoma is 0.34% (90% CrI 0.33– 0.35%) for a person in Southeast Asia uninfected with liver fluke and 4.9% (90% CrI 4.7–5.0%) for a person infected with liver fluke, which is fourteen–fold higher. Our estimate is similar to a previously reported value for Northeast Thailand using registry data from 1998–2002 (43), though we are the first to correct for the proportion of the population exposed to *O. viverrini* by age and to estimate the baseline risk of cholangiocarcinoma in the absence of parasite infection.

We calculate the number of excess cholangiocarcinoma cases in Northeast Thailand attributable to infection with liver fluke by assigning each age group a relative probability of i) baseline risk (*α*_*i*_) and ii) the elevated risk from liver fluke infection given the parasite exposure in that age group (*α*_*i*_ + *βq*_*i*_). Using these probabilities we estimate that 92% of cholangiocarcinoma cases between 1997–2009 are attributable to *O. viverrini* infection (Fig. 3D). A case–control study in Thailand estimated that 91% of cholangiocarcinoma cases in men and 80% in women were attributable to exogenous risk factors (12), which included *O. viverrini* exposure along with betel (areca) nut chewing (known to be an oral carcinogen (44)) and the consumption of certain foods.

Given the availability of data, our estimate of cholangiocarcinoma risk is unstratified by host factors other than age, such as sex, nor is it provided at a more granular spatial scale. The probability of developing cholangiocarcinoma varies with the intensity of liver fluke infection (45) and, given improved longitudinal data, the lifetime probability of cancer should be stratified by the cumulative worm burden in future analyzes.

## Discussion

This study uncovers crucial epidemiological processes on the pathway from parasite infection to malignancy (Table 2), and our objective is for these findings to inform control programs to reduce transmission of liver fluke and subsequent cases of cancer (16, 17). Given our findings that the host age at first fluke infection is before ten years of age and the first amplified driver mutations for biliary cancer occur three decades later, we recommend that interventions should target younger people in endemic regions, such as school–based deworming (46). There is limited evidence on the impact of anthelmintic treatment on preventing or reversing biliary damage in humans (47, 48), although current evidence suggests a beneficial effect of de-worming even in the context of repeated reinfection. Given the dramatic fall in parasite transmission from 1990 in Thailand we predict that the future incidence of cholangiocarcinoma will also decline, though the long temporal lags between initial parasite exposure, carcinogenesis and cancer diagnosis (see Fig. 2B and Fig. 3C) means that changes to *O. viverrini* transmission intensity will take many years to influence the incidence of cholangiocarcinoma at the population level (17). Randomized trials investigating the impact of deworming on the risk of biliary cancer could be considered unethical given the need to restrict anthelmintic treatment for controls (16). Dynamic simulations have a useful role to play, therefore, in estimating the magnitude of interventions over long periods (49). Our results also provide a platform for biomarker discovery by highlighting early driver genes and the long latent period for cholangiocarcinoma provides a window for early therapeutic interventions. Validating predictions of early cholangiocarcinoma driver genes is challenging in humans, although biobanking liver biopsies and bile samples provides an opportunity to prospectively characterize somatic evolution. Ideally, longitudinal studies would record detailed patient information on liver fluke infection and other carcinogenic exposures at multiple time points.

**Table 2.**
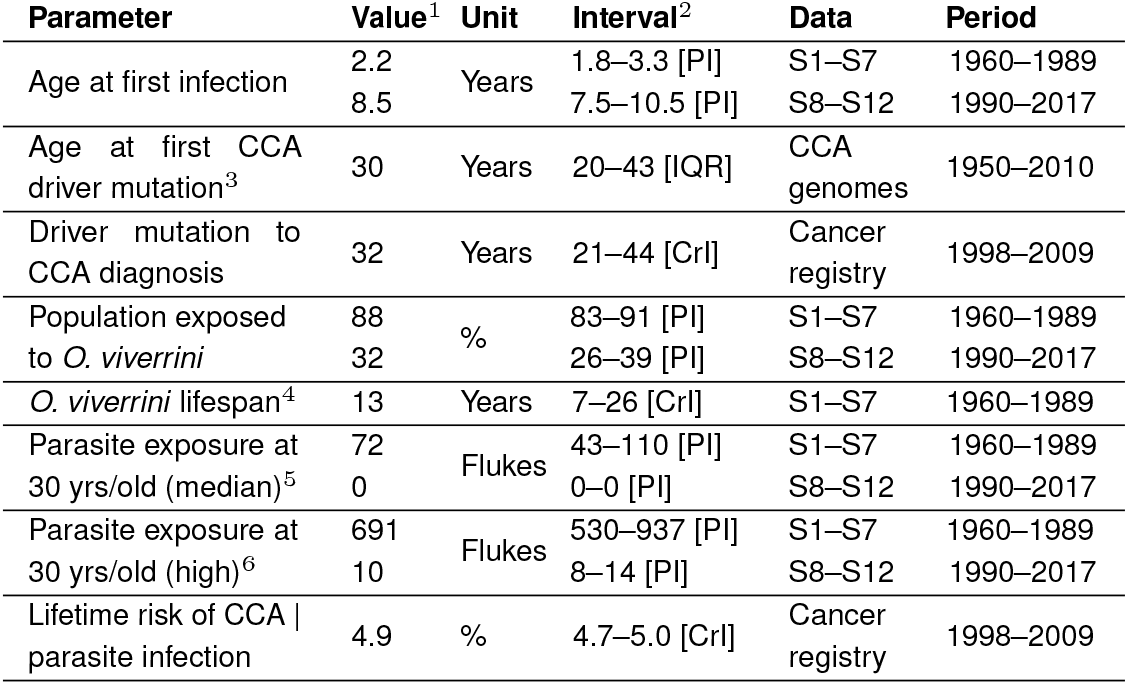
Epidemiological parameters for the liver fluke *Opisthorchis viverrini* and the resulting biliary cancer (cholangiocarcinoma; CCA) estimated in this study using parasitological and cancer genomic data from Thailand. ^1^Posterior median value. ^2^Types of uncertainty intervals are 90% prediction interval [PI], interquartile range [IQR], and 90% credible interval [CrI]. ^3^Age at first amplified driver gene, using C>T mutations at CpG sites under the assumption of a constant mutation rate. ^4^Lifespan of adult *O. viverrini* worms within the human host. ^5^Cumulative parasite burden at 30 years of age given a median exposure (50^*th*^ percentile). ^6^Cumulative parasite burden at 30 years of age given a high exposure (90^*th*^ percentile).

The evolutionary cancer analysis in this study is limited in using single, rather than multiple, tumor and normal sequenced biopsies from each patient, which may result in subclones being under or over represented in the sample (50). The inferred age of amplified mutations is nevertheless similar for clonally and subclonally assigned variants (Fig. 1C), suggesting that our findings are robust. Capturing tumor heterogeneity and clonal inference in future studies would benefit from multi–site sequencing or a single cell approach (51). Novel methods for assigning mutational signatures to SNVs to characterize multiple clock–like signatures and identify mutations specifically associated with parasite exposure would further enhance such studies (25, 52).

Relatively few studies have investigated the epidemiological relationship between infection and carcinogenesis, although compared against other pathogen–driven cancers our estimates of the induction and latent periods for fluke–induced cholangiocarcinoma appear longer. For human papillomavirus (HPV) the time from infection to high-grade cervical intrapithelial neoplasia (CIN2/3) has been estimated from registry data as 3 years, and the time from CIN2/3 to cancer 23.5 years (53). The time from hepatitis B infection to hepatocellular carcinoma is reported as 25–30 years in a review (54) though we are unaware of the supporting evidence. Mechanisms for a potentially longer tumorigenesis from *O. viverrini* infection, compared with other carcinogenic pathogens, have not been investigated. We note that the impact of fluke infection on biliary pathology is dose–dependent (45). There may also be evolutionary reasons for a slower accumulation of pathology from helminths, given their long lifespans (55).

We assume that excess cases of cholangiocarcinoma in North-east Thailand (when compared against Southern Thailand, Malaysia and the Philippines) are solely attributable to liver fluke exposure, though other host factors may be partly responsible. Betel nut chewing and the consumption of fermented foods containing nitrosamines have been implicated as additional risk factors in cross–sectional studies (12, 56). Betal nut chewing occurs in multiple Southeast Asian countries including the Philippines and Malaysia (44), which are used to estimate the baseline risk of cholangiocarcinoma (Figs. 3B and S5). Therefore, our analysis controls for other carcinogenic exposures to the extent that they are practiced throughout Southeast Asia.

As the data used here are primarily from Northeast Thailand our estimates of epidemiological parameters are most applicable to this region. Large–scale ultrasound screening programs for liver disease in Thailand have successfully diagnosed thousands of cholangiocarcinoma and precancerous cases since 2015 (57). However, enhanced screening will likely lead to short–term increases in the reported incidence of cholangiocarcinoma, further complicating our understanding of the relationship between parasite infection and cancer (17). Extending our analyzes to Lao PDR and Cambodia, where the burden of parasitic disease is greater (7, 11) and the capacity for healthcare systems to diagnose and treat biliary cancer more limited (58), remains a priority for future research.

## Methods and Materials

### Cholangiocarcinoma whole–genome sequences

We accessed paired tumor–normal whole–genome sequences from 22 individuals in Northeast Thailand with previous exposure to liver fluke infection. Tumor tissue was obtained from patients during surgical resection of the biliary tract at Srinagarind Hospital in Khon Kaen, Thailand and sequencing was performed on a single core sample per–patient (20). Normal somatic genomes were obtained from patient blood samples. Tumors are classified according to their anatomical location on the biliary tree; namely intrahepatic cholangiocarcinoma (within the hepatic ducts), perihilar cholangiocarcinoma (between the second order bile ducts and the cystic duct insertion) or distal cholangiocarcinoma (below the cystic duct). Here our samples consist of eight perihilar and 14 intrahepatic tumors. The age at surgery ranged from 37–79 years (median 57 years) and 11/22 (50%) of patients were female.

### Cancer Genomics

We trimmed adaptor sequences from 150bp Illumina paired–end reads and mapped the reads against the human reference (GRCh37) using bwa mem v.0.7.17 with default parameters (59). The resulting BAM files were then sorted and indexed with samtools v.1.9. Duplicates were marked and removed using GATK v.4.1.4.1 (60) and base quality scores recalibrated for tumor sequences with ICGC PCAWG consensus calls for somatic single nucleotide variants (SNVs) and indels (61). To examine the depth of mapped reads we took the output from samtools depth -a (including zeroes) and binned the mean depth within 1Mb segments. Sequencing coverage ranged from 35–73x per sample (median 57x) for normal genomes and 45–72x (median 54x) for tumor genomes.

We called SNVs and indels using GATK Mutect2 with a panel of normals provided by the Broad Institute (62) and additional filters to remove secondary and supplementary reads. Before filtering, the number of SNVs ranged from 109,339–292,886 per sample (median 131,057). We calculated the fraction of normal reads with tumor contamination using the GATK tool CalculateContamination in combination with 4.7 million common germline alleles (MAF 0.01–0.20) derived from Asian populations in Singapore (63). This revealed that contamination in normal samples was low with <0.7% of reads coming from cross–sample contamination. Using the contamination data we filtered the variant calls, leaving 2,349–27,821 (median 10,360) SNVs and 268–14,230 (median 1,382) indels per tumor. The number of SNVs called are consistent with other biliary cancers (64).

### Subclonal reconstruction

We estimate tumor copy number using the Battenberg algorithm (65), with reference data from the 1000 Genomes Project. The estimated fraction of tumor cells (rather than normal tissue) in our cholangiocarcinoma genomes, also known as tumor purity or cellularity, varied substantially between samples (range 10–90%; median 60%). The overall ploidy per tumor ranged from 1.2–3.7 (median 2.0). We then phased the somatic variants and assigned them to subclonal lineages (24) using dpclust3p and dpclust (66), implemented in R v.4.3.2. The number of clonal and subclonal lineages, to which variants were assigned, varied from 2–5 per tumor (median 3).

### Timing of driver mutations

We applied algorithms to estimate the chronological timing of driver mutations in amplified regions; MutationTimeR (19) and AmplificationTimeR (21), which were implemented in R v.4.3.3. The MutationTimeR algorithm uses a panel of 371 known driver mutations identified by the PCAWG consortium (64). We used the temporal correlation of copy number gains from MutationTimeR to identify whether tumors had undergone whole–genome duplication (19). For the AmplificationTimeR analysis, we focused on timing the amplification of 43 driver mutations that have been identified as important in early–stage cholangiocarcinoma (22, 23) or previously detected in these tumors (20). Estimates for the chronological time of amplifications were calibrated using C>T mutations at CpG sites (CpG>TpG), which have been established to have clock–like properties and the mutation burden correlates with age (25). The age at driver gene amplification was calculated as the product of the chronological time estimates from AmplificationTimeR (21) in the interval [0, 1] and the patient’s age at surgery (20) under the assumption of a constant CpG>TpG mutation rate.

### Earliest and latest amplified genes

We used a generalized linear model to determine which genes were amplified disproportionately early or late during tumorigenesis. We modeled the chronological time estimates for gene *g* (*y*_*t,g*_), estimated by AmplificationTimeR (21) with a minimum of 10 CpG>TpG mutations, which fall in the interval [0,1] using a beta distribution parameterised by a mean *µ*_*g*_ and precision *κ*

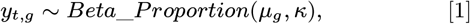

where the mean is a transformed linear function of a gene– specific intercept (*α*_*g*_) plus covariates (*x*) and slopes (*β*). The three binary explanatory variables are the sex of the patient, whether the tumor is intrahepatic or perihilar, and whether the amplification is clonal or subclonal

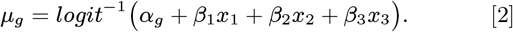

The model was fitted in a Bayesian framework using the stan language v.2.34.1 (67) implemented with cmdstanr v.0.7.1 in R v.4.3.3; see details on parameter estimation below.

### Liver fluke surveys

We obtained data from epidemiological surveys on the liver fluke *O. viverrini* in Thailand between 1980–2017. The parasitological observations in these surveys consist of i) adult worms obtained from liver dissection at autopsy, ii) adult worms recovered through expulsion following anthelminthic treatment, or iii) parasite egg counts obtained by fecal examination, see Table 1. We contacted the study authors to obtain parasitological data by host age. Where these were unavailable, we simulated individual–level data from summary tables which contained the sample size, mean, and variance for parasitological observations (worm burdens or fecal egg counts) aggregated by age group using a Pearson type I distribution.

### Parasite transmission model

We developed a mechanistic model that incorporates key aspects of parasite ecology (26, 68) fitted to individual–level data on adult worm burdens or fecal egg counts (data summarized in Table 1). Our process model characterizes the mean worm burden at the population level (*M*) by host years of age (*a*) as an immigration–death process

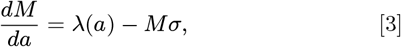

where *σ* gives the spontaneous death rate of adult worms in the absence of anthelmintic treatment. The expected lifespan of adult *O. viverrini* parasites is therefore given as the time taken for half of adult worms to die in the absence of anthelmintic treatment; *log*(2)*/σ*. As there is evidence for age– dependent reinfection rates (69), we model the *O. viverrini* force of infection as a function of host age

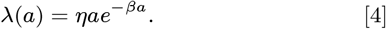

The dynamic model (Equations 3 and 4) has the following analytical solution for worm burden by age *a*,

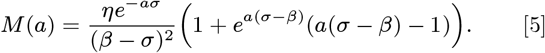

The cumulative parasite exposure (*δ*) by age *a* is given by the definite integral of the force of infection (Equation 4)

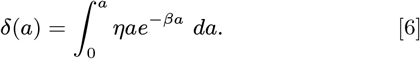

### Observation model for survey data

The true number of *O. viverrini* adult worms per individual *i* of age group *a* (*x*_*i,a*_) follows the negative binomial distribution (*NB*), which takes the form of a gamma–Poisson mixture model parameterized with a mean worm burden *M* (*a*) (Equations 3 and 5) and age– dependent dispersion *k*_*a*_ (37). Values of *k*_*a*_ are estimated for each age–group and are themselves normally distributed with an overall mean of *µ*_*k*_ and a standard deviation of *σ*_*k*_. During autopsy surveys, adult *O. viverrini* were carefully removed from cross–sections of liver and worm recovery is likely close to 100%, therefore we consider that the true worm count for each individual is equal to the recovered worms in autopsy studies (*x*_*i,a*_ ≡ *w*_*i,a*_ | *r* = 1),

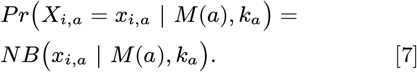

In surveys where *O. viverrini* flukes were obtained by expulsion, participants were treated with the anthelmintic praziquantel followed by a saline purgative, which is known to result in imperfect subsequent recovery of from feces (70). Therefore, we allow the true worm burden to be greater than, or equal to, the observed number of worms for individuals in expulsion studies (*x*_*i,a*_ ≥ *w*_*i,a*_) and the probability of observing *w*_*i*_ worms given a true count of *x*_*i*_ is a binomial sampling process with probability of worm recovery *r* = 0.44 (37),

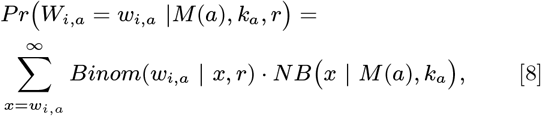

where *Binom* refers to a standard binomial probability distribution. In surveys where the outcome variables are eggs per gram of stool (*y*), if at least one egg is observed for individual *i* of age group *a* (*y*_*i,a*_ ≥1), we relate this to the expected egg count for that individual using a negative binomial error distribution, where the mean is given as a density–dependent function of the true worm burden; *π*(*x*) = (Λ*x*)^*γ*^ and the dispersion is given by parameter *h*, which has previously been estimated for *O. viverrini* as 0.4 (37),

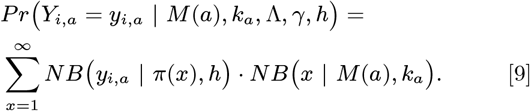

Where zero eggs are observed (*y*_*i,a*_ = 0), we consider the individual diagnostic sensitivity as a saturating function of the worm burden, *se*(*x*) = *x/*(*x* + *b*), where the parameter *b* has been previously estimated for *O. viverrini* as 1.7 (37),

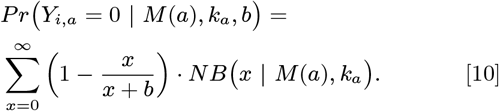

The population level sensitivity (*S*) for fecal egg diagnostics is a function of the worm burden distribution at the population level (37),

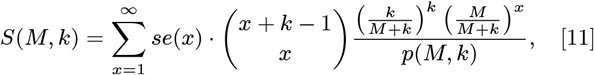

where *p*(*M, k*) indicates the true prevalence and is given by

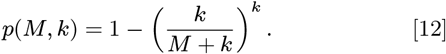

Given an assumed diagnostic specificity of one, the observed prevalence *p*^*′*^ for for age group *a* is therefore related to the true prevalence with the following expression,

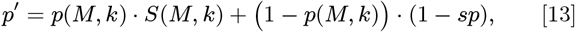

where *sp* gives the fecal egg diagnostic specificity, which is assumed here to be 1. For surveys where only the prevalence is given by fecal egg diagnostic, we represent this as individual– level positive or negative outcomes; *z*_*i,a*_ ∈{0, 1}. The probability for the binary diagnostic observations is therefore given by a Bernoulli distribution

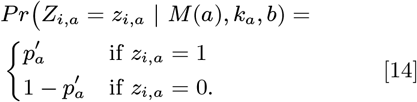

### Epidemiological parameter estimation

Model fitting was performed in a Bayesian framework using the stan language v.2.34.1 (67) implemented with cmdstanr v.0.7.1 (71) in R v.4.3.3. Parameters were assigned weakly or moderately informative prior distributions based on the results from a previous analysis (37) for the pre–intervention data (S1–S7, see Table 1). For the post–intervention analysis (S8–S12), several model parameter values were taken directly from the pre–intervention posterior distributions; including the parameters relating worm burdens to egg counts (Λ, *γ*) and the worm mortality rate (*σ*), which was taken as the highest percentile of the posterior estimate (*σ* = 0.116), corresponding to a mean worm lifespan of 6 years, to account for higher parasite mortality resulting from periodic anthelmintic treatment after 1990. Each model was run with four parallel chains with a burn-in of 1700 iterations per chain and a total of 1000 iterations. The Gelman–Rubin diagnostic 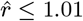 and effective sample size > 500 were used to diagnose successful Markov chain convergence. Results are presented as credible intervals (CrI) of parameter posterior distribution or prediction intervals (PI) from simulations.

### Induction and latent period

We performed a time–to–event analysis to validate the induction and latent periods estimated in the evolutionary cancer analysis using cancer registry data. We obtained the age distribution of 10,737 cholangiocarcinoma cases diagnosed at Srinagarind Hospital between 1985–2009 (40) that were grouped into age classes. We consider that for each cancer case at age *a* there is an unknown age at which a driver mutation was obtained *m*, where *m < a*. Values of *m* are drawn from a gamma distribution with mean *µ*_*mut*_ and shape parameter *α*_*mut*_. Following the mutation at age *m* there is a latent period before cancer diagnosis at age *a*, this latent period is also gamma distributed with mean *µ*_*cca*_ and shape *α*_*cca*_. The probability of developing cholangiocarcinoma at age *a, ϕ*_*a*_ is therefore

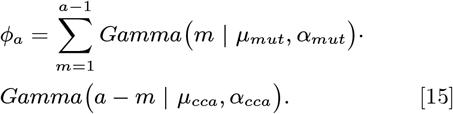

### Probability of acquiring cholangiocarcinoma

We adopt a binomial regression framework with the probability of contracting cholangiocarcinoma in age group *i*, population *j* given by

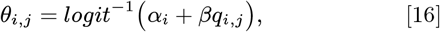

where *α* is an intercept which varies by age group *i* and *β* is a coefficient multiplied by the proportion of the population exposed to liver fluke *q*. For the population in Northeast Thailand, *q*_*i,j*_ is taken as the pre–intervention prevalence for age groups ≥ 20 years old and the post–intervention prevalence for age groups <20 years old. In non–fluke populations (Southern Thailand, Malaysia, and the Philippines) *q*_*i,j*_ = 0. Counts of cholangiocarcinoma cases *c*_*i,j*_ are reported by age group *i* with an underlying population size of *N*_*i,j*_. The likelihood is therefore given by

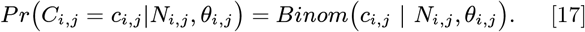

To calculate the lifetime risk of cholangiocarcinoma under the counterfactual scenarios of either infection with liver fluke (*q* = 1) or uninfected (*q* = 0), we obtain the adjusted unconditional probability of cholangiocarcinoma by summing the probabilities in each five year age interval *i* ∈ {1, 2, …, *m*}

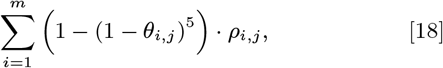

where *ρ*_*i,j*_ is the probability of survival to age group *i* given by demographic life tables for Thailand and Malaysia (72).

For the fluke–endemic population in the survival analysis, we used counts of cholangiocarcinoma cases from the cancer registry at Srinagarind Hospital in Khon Kaen (40) between 1998–2009 along with corresponding demographic data (*N*) in the same period (73). Cholangiocarcinoma case data from Southern Thailand (Songkla province) between 2004–2013 was used to calculate the baseline risk (74), along with cancer and demographic data for Malaysia (2007–2009) and the Philippines (2008) obtained from the World Health Organization Mortality Database using version 10 ICD codes C22 (malignant neoplasm of liver and intrahepatic bile ducts) and C24 (malignant neoplasm of other and unspecified parts of biliary tract) (75). We assumed that half the biliary cancers in Malaysia and the Philippines were attributable to cholangiocarcinoma (14) and that, given the very low survival rate, mortality is a valid approximation for incidence (15).

## Data Availability

Code and publicly available data to reproduce the analysis are available at https://github.com/tc13/ov-cca-models.
Access to sequence data on biliary cancer genomes is controlled by the International Cancer Genome Consortium Data Access Compliance Office (ICGC DACO).

## Acknowledgements

We thank Dr. Stefan Dentro and Prof. Moritz Gerstung for providing additional code relating to the PCAWG consortium analysis to calculate mutation times of 2,778 tumor genomes. We appreciate the comments from Dr. Arporn Wangwiwatsin and Prof. Watcharin Loilome on a poster relating to this manuscript. We thank Prof. Simon E.F. Spencer for discussions on the survival analysis. Dr. Claudio Nunes–Alves and Ms. Hannah R. McNeill also provided valuable feedback on the manuscript.

Access to whole-genome paired cancer and normal sequence data was approved by the International Cancer Genome Consortium Data Access Compliance Office (reference: DACO-63). We acknowledge the important role of Prof. Apinya Jusakul, Prof. Patrick Tan and colleagues for generating this sequence data.

Access to the SG10K_Pilot data was approved by the National Precision Medicine Data Access Committee in Singapore (Application no. SG10KP00043). We thank the ‘SG10K_Pilot Investigators’ for providing the ‘SG10K_Pilot data’ (EGAD00001005337). The data from the ‘SG10K_Pilot Study’ reported here were obtained from EGA. This manuscript was not prepared in collaboration with the ‘SG10K_Pilot Study’ and does not necessarily reflect the opinions or views of the ‘SG10K_Pilot Study’.

We used data from the World Health Organization (WHO) Mortality Database for biliary cancer incidence. The analyses, interpretations and conclusions shown in this article are attributable solely to the authors and not to WHO, which is responsible only for the provision of the original information.

This research was funded in whole, or in part, by the Wellcome Trust [Grant number 215919/B/19/Z]. For the purpose of open access, the author has applied a CC BY public copyright licence to any Author Accepted Manuscript version arising from this submission.

C.B. acknowledges that this study was performed under the collaborative umbrella of the European Network for the Study of Cholangiocarcinoma (EURO-CHOLANGIO-NET) supported by COST action (CA) 18122 and the Precision-BTC network supported by CA22125. F.V. acknowledges funding from a short term grant from EURO-CHOLANGIO-NET CA18122.

P.S. acknowledges funding from the National Research Council of Thailand as part of the Fluke–Free Thailand program.

## Ethical Statement

As an analysis of previously published and anonymous human data, this study met the criteria for exemption from ethical review at the Universities of Oxford and Glasgow.

## Data and Code Accessibility

This study uses previously published datasets. Code to reproduce the epidemiological analysis is available at https://github.com/tc13/ov-cca-models/. Cholangiocarcinoma whole–genomes were accessed from the European Genome–Phenome archive under accessions EGAD00001001988 and EGAD00001003834.

## Authors’ Contributions

T.C. conceptualized the study, T.C. developed methodology, T.C. analyzed epidemiological data, T.C and analyzed cancer genomic data, P.S provided resources, C.B. provided supervision for cancer genomics, T.D.H. provided supervision for epidemiological modeling, T.C. wrote the manuscript.

## Footnotes

* https://monographs.iarc.fr/wp-content/uploads/2018/06/mono61.pdf

† https://monographs.iarc.fr/wp-content/uploads/2018/06/mono100B.pdf

## References

1. Catherine de Martel, Damien Georges, Freddie Bray, Jacques Ferlay, and Gary M Clifford. Global burden of cancer attributable to infections in 2018: a worldwide incidence analysis. The Lancet global health, 8(2):e180–e190, 2020.

2. Anna Borlase, Joaquin M Prada, and Thomas Crellen. Modelling morbidity for neglected tropical diseases: the long and winding road from cumulative exposure to long-term pathology. Philosophical Transactions of the Royal Society B, 378(1887):20220279, 2023.

3. Laila Sara Arroyo Mühr, Andrea Gini, Emel Yilmaz, Sadaf S Hassan, Camilla Lagheden, Emilie Hultin, Ainhoa Garcia Serrano, Agustin E Ure, Helena Andersson, Roxana Merino, et al. Concomitant human papillomavirus (hpv) vaccination and screening for elimination of hpv and cervical cancer. Nature Communications, 15(1):3679, 2024.

4. Vanessa O Ezenwa and Anna E Jolles. Opposite effects of anthelmintic treatment on microbial infection at individual versus population scales. Science, 347(6218):175–177, 2015.

5. Sergi Alonso, Moses Arinaitwe, Alon Atuhaire, Andrina Barungi Nankasi, Joaquín M Prada, Emma McIntosh, and Poppy HL Lamberton. The short-term impact of schistosoma mansoni infection on health-related quality of life: implications for current elimination policies. Proceedings B, 291(2024):20240449, 2024.

6. Carl Grundy-Warr, Ross H Andrews, Paiboon Sithithaworn, Trevor N Petney, Banchop Sripa, Luxana Laithavewat, and Alan D Ziegler. Raw attitudes, wetland cultures, life-cycles: socio-cultural dynamics relating to opisthorchis viverrini in the mekong basin. Parasitology international, 61(1):65–70, 2012.

7. Ting-Ting Zhao, Yi-Jing Feng, Pham Ngoc Doanh, Somphou Sayasone, Virak Khieu, Choosak Nithikathkul, Men-Bao Qian, Yuan-Tao Hao, and Ying-Si Lai. Model-based spatial-temporal mapping of opisthorchiasis in endemic countries of southeast asia. elife, 10:e59755, 2021.

8. P Jongsuksuntigul and T Imsomboon. The impact of a decade long opisthorchiasis control program in northeastern thailand. The Southeast Asian Journal of Tropical Medicine and Public Health, 28(3):551–557, 1997.

9. Apiporn Suwannatrai, Prasert Saichua, and Melissa Haswell. Epidemiology of Opisthorchis viverrini infection. Advances in Parasitology, 101:41–67, 2018.

10. Oranard Wattanawong, Sopon Iamsirithaworn, Thongroo Kophachon, Worayuth Nak-Ai, Ampas Wisetmora, Thitima Wongsaroj, Paron Dekumyoy, Choosak Nithikathkul, Apiporn T Suwannatrai, and Banchob Sripa. Current status of helminthiases in Thailand: A cross-sectional, nationwide survey, 2019. Acta Tropica, 223:106082, 2021.

11. Virak Khieu, Thomas Fürst, Kazuko Miyamoto, Tai-Soon Yong, Jong-Yil Chai, Rekol Huy, Sinuon Muth, and Peter Odermatt. Is opisthorchis viverrini emerging in cambodia? Advances in parasitology, 103:31–73, 2019.

12. D Maxwell Parkin, Petcharin Srivatanakul, Myriam Khlat, Dhiraphol Chenvidhya, Pornarong Chotiwan, Somchai Insiripong, Kristan A L’abbé, and Christopher P Wild. Liver cancer in thailand. i. a case-control study of cholangiocarcinoma. International journal of cancer, 48(3): 323–328, 1991.

13. Andrea A Florio, Jacques Ferlay, Ariana Znaor, David Ruggieri, Christian S Alvarez, Mathieu Laversanne, Freddie Bray, Katherine A McGlynn, and Jessica L Petrick. Global incidence and trends in intra-and extrahepatic cholangiocarcinoma from 1993 to 2012. Cancer, 126(11):2666, 2020.

14. Banchob Sripa, Paul J Brindley, Jason Mulvenna, Thewarach Laha, Michael J Smout, Eimorn Mairiang, Jeffrey M Bethony, and Alex Loukas. The tumorigenic liver fluke Opisthorchis viverrini–multiple pathways to cancer. Trends in Parasitology, 28(10):395–407, 2012.

15. Sumera Rizvi, Shahid A Khan, Christopher L Hallemeier, Robin K Kelley, and Gregory J Gores. Cholangiocarcinoma—evolving concepts and therapeutic strategies. Nature reviews Clinical oncology, 15(2):95–111, 2018.

16. Thomas Crellen, Paiboon Sithithaworn, Opal Pitaksakulrat, Narong Khuntikeo, Graham F Medley, and T Déirdre Hollingsworth. Towards evidence-based control of Opisthorchis viverrini. Trends in Parasitology, 37(5):370–380, 2021.

17. Narong Khuntikeo, Bandit Thinkhamrop, Thomas Crellen, Chatanun Eamudomkarn, Trevor N Petney, Ross H Andrews, and Paiboon Sithithaworn. Epidemiology and control of opisthorchis viverrini infection: Implications for cholangiocarcinoma prevention. In Liver Fluke, Opisthorchis viverrini Related Cholangiocarcinoma: Liver Fluke Related Cholangiocarcinoma, pages 27–52. Springer, 2023.

18. Andrea Sottoriva, Inmaculada Spiteri, Sara GM Piccirillo, Anestis Touloumis, V Peter Collins, John C Marioni, Christina Curtis, Colin Watts, and Simon Tavaré. Intratumor heterogeneity in human glioblastoma reflects cancer evolutionary dynamics. Proceedings of the National Academy of Sciences, 110(10):4009–4014, 2013.

19. Moritz Gerstung, Clemency Jolly, Ignaty Leshchiner, Stefan C Dentro, Santiago Gonzalez, Daniel Rosebrock, Thomas J Mitchell, Yulia Rubanova, Pavana Anur, Kaixian Yu, et al. The evolutionary history of 2,658 cancers. Nature, 578(7793):122–128, 2020.

20. Apinya Jusakul, Ioana Cutcutache, Chern Han Yong, Jing Quan Lim, Mi Ni Huang, Nisha Padmanabhan, Vishwa Nellore, Sarinya Kongpetch, Alvin Wei Tian Ng, Ley Moy Ng, et al. Whole-genome and epigenomic landscapes of etiologically distinct subtypes of cholangiocarcinoma. Cancer discovery, 7(10):1116–1135, 2017.

21. G Maria Jakobsdottir, Stefan C Dentro, Robert G Bristow, and David C Wedge. Amplification-timer: An r package for timing sequential amplification events. Bioinformatics, page btae281, 2024.

22. Benjamin Goeppert, Damian Stichel, Reka Toth, Sarah Fritzsche, Moritz Anton Loeffler, Anna Melissa Schlitter, Olaf Neumann, Yassen Assenov, Monika Nadja Vogel, Arianeb Mehrabi, et al. Integrative analysis reveals early and distinct genetic and epigenetic changes in intraductal papillary and tubulopapillary cholangiocarcinogenesis. Gut, 71(2):391–401, 2022.

23. Khaa Hoo Ong, Yao-Yu Hsieh, Hong-Yue Lai, Ding-Ping Sun, Tzu-Ju Chen, STEvEN KuAN-HuA HuANG, Yu-Feng Tian, Chia-Lin Chou, Yow-Ling Shiue, Hung-Chang Wu, et al. Lamc2 is a potential prognostic biomarker for cholangiocarcinoma. Oncology Letters, 26(6):1–14, 2023.

24. Stefan C Dentro, David C Wedge, and Peter Van Loo. Principles of reconstructing the subclonal architecture of cancers. Cold Spring Harbor perspectives in medicine, 7(8):a026625, 2017.

25. Ludmil B Alexandrov, Jaegil Kim, Nicholas J Haradhvala, Mi Ni Huang, Alvin Wei Tian Ng, Yang Wu, Arnoud Boot, Kyle R Covington, Dmitry A Gordenin, Erik N Bergstrom, et al. The repertoire of mutational signatures in human cancer. Nature, 578(7793):94–101, 2020.

26. GF Medley and DA Bundy. Dynamic modeling of epidemiologic patterns of schistosomiasis morbidity. The American journal of tropical medicine and hygiene, 55(5 Suppl):149–158, 1996.

27. Paiboon Sithithaworn, Smarn Tesana, Vichit Pipitgool, Sasithorn Kaewkes, Kasemsri Thaiklar, Chavalit Pairojkul, Bunchob Sripa, Anucha Paupairoj, Pisit Sanpitak, and Chanarong Aranyanat. Quantitative post-mortem study of Opisthorchis viverrini in man in North-East Thailand. Transactions of the Royal Society of Tropical Medicine and Hygiene, 85(6):765–768, 1991.

28. Roger J Ramsay, Paiboon Sithithaworn, Paul Prociv, Douglas E Moorhouse, and Chingchai Methaphat. Density-dependent fecundity of Opisthorchis viverrini in humans, based on faecal recovery of flukes. Transactions of the Royal Society of Tropical Medicine and Hygiene, 83(2): 241–242, 1989.

29. MR Haswell-Elkins, DB Elkins, Paiboon Sithithaworn, Phattara Treesarawat, and Sasithorn Kaewkes. Distribution patterns of opisthorchis viverrini within a human community. Parasitology, 103(1):97–101, 1991.

30. ES Upatham, V Viyanant, S Kurathong, J Rojborwonwitaya, WY Brockelman, S Ardsungnoen, P Lee, and S Vajrasthira. Relationship between prevalence and intensity of opisthorchis viverrini infection, and clinical symptoms and signs in a rural community in north-east thailand. Bulletin of the World Health Organization, 62(3):451, 1984.

31. Sucha Kurathong, Pravit Lerdverasirikul, Virasak Wongpaitoon, Chutima Pramoolsinsap, and E Suchart Upatham. Opisthorchis viverrini infection in rural and urban communities in northeast thailand. Transactions of the Royal Society of Tropical Medicine and Hygiene, 81(3):411–414, 1987.

32. S Sornmani, FP Schelp, P Vivatanasesth, W Patihatakorn, P Impand, P Sitabutra, P Worasan, and S Preuksaraj. A pilot project for controlling o. viverrini infection in nong wai, northeast thailand, by applying praziquantel and other measures. Arzneimittel-forschung, 34(9B):1231–1234, 1984.

33. Soraya J Kaewpitoon, Ratana Rujirakul, and Natthawut Kaewpitoon. Prevalence of opisthorchis viverrini infection in nakhon ratchasima province, northeast thailand. Asian Pacific Journal of Cancer Prevention, 13(10):5245–5249, 2012.

34. Kesorn Thaewnongiew, Seri Singthong, Saowalux Kutchamart, Sasithorn Tangsawad, Supannee Promthet, Supan Sailugkum, and Narong Wongba. Prevalence and risk factors for opisthorchis viverrini infections in upper northeast thailand. Asian Pacific Journal of Cancer Prevention, 15(16):6609–6612, 2014.

35. Pokkamol Laoraksawong, Oranuch Sanpool, Rutchanee Rodpai, Tongjit Thanchomnang, Wanida Kanarkard, Wanchai Maleewong, Ratthaphol Kraiklang, and Pewpan M Intapan. Current high prevalences of strongyloides stercoralis and opisthorchis viverrini infections in rural communities in northeast thailand and associated risk factors. BMC Public Health, 18:1–11, 2018.

36. DJ Shaw, BT Grenfell, and AP Dobson. Patterns of macroparasite aggregation in wildlife host populations. Parasitology, 117(6):597–610, 1998.

37. Thomas Crellen, Melissa Haswell, Paiboon Sithithaworn, Somphou Sayasone, Peter Odermatt, Poppy HL Lamberton, Simon EF Spencer, and T Déirdre Hollingsworth. Diagnosis of helminths depends on worm fecundity and the distribution of parasites within hosts. Proceedings of the Royal Society B, 290(1991):20222204, 2023.

38. Mark Viney and Jo Cable. Macroparasite life histories. Current Biology, 21(18):R767–R774, 2011.

39. HD Attwood and ST Chou. The longevity of Clonorchis sinensis. Pathology, 10(2):153–156, 1978.

40. Supot Kamsa-ard, Surapon Wiangnon, Krittika Suwanrungruang, Supannee Promthet, Narong Khuntikeo, Siriporn Kamsa-ard, Suwannee Mahaweerawat, et al. Trends in liver cancer incidence between 1985 and 2009, khon kaen, thailand: cholangiocarcinoma. Asian Pac J Cancer Prev, 12(9):2209–2213, 2011.

41. George Psevdos, Florence M Ford, and Sung-Tae Hong. Screening US vietnam veterans for liver fluke exposure 5 decades after the end of the war. Infectious Diseases in Clinical Practice, 26(4):208–210, 2018.

42. Theodore E Nash, David Sullivan, Edward Mitre, Eric Garges, Victoria J Davey, Gary Roselle, Stephen J Davies, and Peter D Rumm. Comments on “Screening US Vietnam veterans for liver fluke exposure 5 decades after the end of the war”. Infectious Diseases in Clinical Practice, 26 (4):240–241, 2018.

43. Nopparat Songserm, Supannee Promthet, Paiboon Sithithaworn, Chamsai Pientong, Tipaya Ekalaksananan, Peechanika Chopjitt, and Donald Maxwell Parkin. Risk factors for cholangiocarcinoma in high-risk area of thailand: role of lifestyle, diet and methylenetetrahydrofolate reductase polymorphisms. Cancer epidemiology, 36(2):e89–e94, 2012.

44. Saman Warnakulasuriya, Chetan Trivedy, and Timothy J Peters. Areca nut use: an independent risk factor for oral cancer: The health problem is under-recognised. BMJ, 324(7341):799–800, 2002.

45. Melissa R Haswell-Elkins, Soisungwan Satarug, Mitsuhiro Tsuda, Eimorn Mairiang, Hiroyasu Esumi, Paiboon Sithithaworn, Pisain Mairiang, Minoru Saitoh, Puangrat Yongvanit, and David B Elkins. Liver fluke infection and cholangiocarcinoma: model of endogenous nitric oxide and extragastric nitrosation in human carcinogenesis. Mutation Research/Fundamental and Molecular Mechanisms of Mutagenesis, 305(2):241–252, 1994.

46. Joanne P Webster, David H Molyneux, Peter J Hotez, and Alan Fenwick. The contribution of mass drug administration to global health: past, present and future. Philosophical Transactions of the Royal Society B: Biological Sciences, 369(1645):20130434, 2014.

47. Eimorn Mairiang, Melissa R Haswell-Elkins, Pisaln Mairiang, Paiboon Sithithaworn, and David B Elkins. Reversal of biliary tract abnormalities associated with Opisthorchis viverrini infection following praziquantel treatment. Transactions of the Royal Society of Tropical Medicine and Hygiene, 87(2):194–197, 1993.

48. S Pungpak, C Viravan, B Radomyos, K Chalermrut, C Yemput, W Plooksawasdi, M Ho, T Harinasuta, and D Bunnag. Opisthorchis viverrini infection in thailand: studies on the morbidity of the infection and resolution following praziquantel treatment. The American journal of tropical medicine and hygiene, 56(3):311–314, 1997.

49. Sue J Goldie, Daniel Grima, Michele Kohli, Thomas C Wright, Milton Weinstein, and Eduardo Franco. A comprehensive natural history model of hpv infection and cervical cancer to estimate the clinical impact of a prophylactic hpv-16/18 vaccine. International Journal of cancer, 106(6): 896–904, 2003.

50. James RM Black and Nicholas McGranahan. Genetic and non-genetic clonal diversity in cancer evolution. Nature Reviews Cancer, 21(6):379–392, 2021.

51. Zaira Seferbekova, Artem Lomakin, Lucy R Yates, and Moritz Gerstung. Spatial biology of cancer evolution. Nature Reviews Genetics, 24(5):295–313, 2023.

52. Hu Jin, Doga C Gulhan, Benedikt Geiger, Daniel Ben-Isvy, David Geng, Viktor Ljungström, and Peter J Park. Accurate and sensitive mutational signature analysis with musical. Nature Genetics, 56(3):541–552, 2024.

53. Margaretha A Vink, Johannes A Bogaards, Folkert J van Kemenade, Hester E de Melker, Chris JLM Meijer, and Johannes Berkhof. Clinical progression of high-grade cervical intraepithelial neoplasia: estimating the time to preclinical cervical cancer from doubly censored national registry data. American journal of epidemiology, 178(7):1161–1169, 2013.

54. William M Lee. Hepatitis b virus infection. New England journal of medicine, 337(24):1733–1745, 1997.

55. SA Frank and P Schmid-Hempel. Mechanisms of pathogenesis and the evolution of parasite virulence. Journal of evolutionary biology, 21(2):396–404, 2008.

56. P Srivatanakul, S Sukaryodhin, H Ohshima, M Khlat, M Parkin, I Brouet, and H Bartsch. Opisthorchis viverrini infestation and endogenous nitrosamines as risk factors for cholangiocarcinoma in thailand. International journal of cancer, 48(6):821–825, 1991.

57. Kavin Thinkhamrop, Narong Khuntikeo, Nittaya Chamadol, Apiporn T Suwannatrai, Surachai Phimha, and Matthew Kelly. Associations between ultrasound screening findings and cholangiocarcinoma diagnosis in an at-risk population. Scientific Reports, 12(1):13513, 2022.

58. Phonepasong Ayé Soukhathammavong, Virasack Rajpho, Khampheng Phongluxa, Youthanavanh Vonghachack, Jan Hattendorf, Bouasy Hongvanthong, Oroth Rasaphon, Banchob Sripa, Kongsap Akkhavong, Christoph Hatz, et al. Subtle to severe hepatobiliary morbidity in opisthorchis viverrini endemic settings in southern laos. Acta tropica, 141:303–309, 2015.

59. Heng Li and Richard Durbin. Fast and accurate short read alignment with burrows–wheeler transform. bioinformatics, 25(14):1754–1760, 2009.

60. Geraldine A Van der Auwera and Brian D O’Connor. Genomics in the cloud: using Docker, GATK, and WDL in Terra. O’Reilly Media, 2020.

61. PCAWG. URL https://dcc.icgc.org/releases/PCAWG/consensus_snv_indel. DACO.

62. Broad Institute. URL https://console.cloud.google.com/storage/browser/gatk-best-practices. GATK Best Practices.

63. Degang Wu, Jinzhuang Dou, Xiaoran Chai, Claire Bellis, Andreas Wilm, Chih Chuan Shih, Wendy Wei Jia Soon, Nicolas Bertin, Clarabelle Bitong Lin, Chiea Chuen Khor, et al. Large-scale whole-genome sequencing of three diverse asian populations in singapore. Cell, 179(3): 736–749, 2019.

64. The ICGC/TCGA Pan-Cancer Analysis of Whole Genomes Consortium. Pan-cancer analysis of whole genomes. Nature, 578(7793):82–93, 2020.

65. Serena Nik-Zainal, Peter Van Loo, David C Wedge, Ludmil B Alexandrov, Christopher D Greenman, King Wai Lau, Keiran Raine, David Jones, John Marshall, Manasa Ramakrishna, et al. The life history of 21 breast cancers. Cell, 149(5):994–1007, 2012.

66. Niccolo Bolli, Hervé Avet-Loiseau, David C Wedge, Peter Van Loo, Ludmil B Alexandrov, Inigo Martincorena, Kevin J Dawson, Francesco Iorio, Serena Nik-Zainal, Graham R Bignell, et al. Heterogeneity of genomic evolution and mutational profiles in multiple myeloma. Nature communications, 5(1):2997, 2014.

67. Bob Carpenter, Andrew Gelman, Matthew D Hoffman, Daniel Lee, Ben Goodrich, Michael Betancourt, Marcus Brubaker, Jiqiang Guo, Peter Li, and Allen Riddell. Stan: A probabilistic programming language. Journal of Statistical Software, 76(1), 2017.

68. Anna Borlase, James W Rudge, Elsa Léger, Nicolas D Diouf, Cheikh B Fall, Samba D Diop, Stefano Catalano, Mariama Sène, and Joanne P Webster. Spillover, hybridization, and persistence in schistosome transmission dynamics at the human–animal interface. Proceedings of the National Academy of Sciences, 118(41), 2021.

69. ES Upatham, V Viyanant, WY Brockelman, S Kurathong, P Lee, and R Kraengraeng. Rate of reinfection by Opisthorchis viverrini in an endemic northeast Thai community after chemotherapy. International Journal for Parasitology, 18(5):643–649, 1988.

70. Chenghua Shen, Jae-hwan Kim, Jeong-Keun Lee, Young Mee Bae, Min-Ho Choi, Jin-Kyoung Oh, Min Kyung Lim, Hai-Rim Shin, and Sung-Tae Hong. Collection of Clonorchis sinensis adult worms from infected humans after praziquantel treatment. The Korean Journal of Parasitology, 45(2):149, 2007.

71. Jonah Gabry, Rok Češnovar, and Andrew Johnson. cmdstanr: R Interface to ‘CmdStan’, 2023. https://mc-stan.org/cmdstanr/, https://discourse.mc-stan.org.

72. Max Planck Institute for Demographic Research (Germany), University of California, Berkeley (USA), and French Institute for Demographic Studies (France). Human Mortality Database. Available at http://www.lifetable.de (data downloaded on 11th June 2025).

73. Thomas Brinkhoff. website, February 2024. URL http://www.citypopulation.de. City Population.

74. Seesai Yeesoonsang, Edward McNeil, Shama Virani, Surichai Bilheem, Chakrarat Pittayawonganon, Chuleeporn Jiraphongsa, and Hutcha Sriplung. Trends in Incidence of Two Major Subtypes of Liver and Bile Duct Cancer: Hepatocellular Carcinoma and Cholangiocarcinoma in Songkhla, Southern Thailand, 1989–2030. Journal of Cancer Epidemiology, 2018(1):8267059, 2018.

75. World Health Organization. website, February 2024. URL https://www.who.int/data/data-collection-tools/who-mortality-database. WHO Mortality Database.

